# *BCKDK* rs14235 A allele is associated with milder motor impairment and altered network topology in Parkinson’s disease

**DOI:** 10.1101/2023.07.20.23292985

**Authors:** Zhichun Chen, Bin Wu, Guanglu Li, Liche Zhou, Lina Zhang, Jun Liu

## Abstract

**Background:** A multitude of genetic variants confer a risk of Parkinson’s disease (PD), however, whether these risk variants affected the motor symptoms of PD patients remain largely elusive. The objective of this study is to investigate the effects of *BCKDK* rs14235 (G > A), a risk variant associated with PD risk, on the motor manifestations and brain networks of PD patients.

**Methods:** PD patients (n = 146) receiving magnetic resonance imaging from Parkinson’s Progression Markers Initiative (PPMI) database were investigated. The effects of *BCKDK* rs14235 on the motor manifestations and brain networks of PD patients were systematically evaluated.

**Results:** *BCKDK* rs14235 A allele was associated with milder motor symptoms in PD patients. *BCKDK* rs14235 significantly modified the topology of brain structural and functional network. The assortativity in structural network was negatively associated with rigidity in PD while the shortest path length of right pallidum was positively associated with Unified Parkinson’s Disease Rating Scale part III (UPDRS-III) scores. The mediation analysis suggested that assortativity in structural network mediated the effects of *BCKDK* rs14235 on rigidity and the shortest path length of right pallidum mediated the effects of *BCKDK* rs14235 on UPDRS-III scores.

**Conclusions:** *BCKDK* rs14235 significantly shaped the motor impairment and network topology of PD patients. Differential network metrics mediated the effects of *BCKDK* rs14235 on rigidity and UPDRS-III scores of PD patients. Future studies were required to identify the molecular mechanisms underlying the effects of *BCKDK* rs14235 on motor impairment and brain network metrics of PD patients.

## Introduction

Parkinson’s disease (PD) is the most common movement disorder in the world ^1^. Bradykinesia, rigidity, static tremor, postural impairment, and gait disabilities were the typical motor manifestations in PD patients ^1^. Although the loss of dopaminergic neurons was generally thought to be the major culprit resulting in motor impairment in PD, the mechanisms underlying the degeneration of dopaminergic neurons remain largely elusive ^1^. According to previous literature, both environmental and genetic factors contribute to dopaminergic neurodegeneration in PD ^2–8^, however, the occurrence of PD in most individuals is considered to be idiopathic with an unknown cause. Currently, hundreds of genetic loci conferring a risk of PD have been identified^5,6^, nevertheless, the neural mechanisms underlying the pathophysiology of these risk loci for PD are mainly unknown. Decoding the pathophysiology of PD from the perspective of genetics has recently become the mainstream study area as a result of the identification of new risk loci ^5–7^. Recently, new research breakthroughs in PD have been driven by the discovery of the biological functions of PD risk genes and their roles in the occurrence and development of PD at molecular, cellular, neural circuit and behavior levels ^9–16^. Over the last decade, a number of genes associated with PD risk, including *SNCA*^14,17–19^, *PRKN*^9–12^, *LRRK2*^20–25^, *PINK1* ^11–13^, *TMEM175* ^16^, *GPNMB* ^26,27^, and *ATP13A2* ^15^, have been studied, both in terms of physiological and pathological functions. These discoveries have expanded our understanding of the neurodegenerative process in PD and identified key mechanisms by which we can identify potential therapeutic targets.

There is mounting evidence that the clinical characteristics of individuals with PD were diverse and intricate, presenting a significant obstacle to the clinical diagnosis and management in PD patients ^28–31^. Previous studies suggest that genetic variations modify age at onset ^32,33^, influence motor progression ^34–36^, and alter the frequency of a variety of nonmotor symptoms ^37–39^. Specifically, a multitude of mutated genes, such as *GBA* ^36,38,39^, *LRRK2* ^24,40,41^, *PRKN* ^42,43^, *PINK1* ^43,44^, *SNCA* ^41,45^ have been proven to alter the diverse range of symptoms experienced by individuals with familial PD. Therefore, genetic variations significantly contributed to the clinical heterogeneity of PD patients. For example, it has been shown that individuals harboring *GBA* variants encountered an earlier age at onset and exhibited more pronounced motor and nonmotor symptoms in comparison to individuals without GBA variants ^46,47^. According to previous studies, the motor impairment of PD patients were linked to several variants, including *GBA* ^46,47^, *SNCA* ^48,49^, *BST1* ^50^, *ATP8B2* ^51^, *PARK16* ^52^, *GCH1* ^53^, and *APOE* ^54^. Recently, we revealed 3 PD-risk associated genetic variants, *BCKDK* rs14235, *CCDC62* rs11060180, and *GCH1* rs11158026, were significantly associated with motor function, which was measured by Unified Parkinson’s Disease Rating Scale part III (UPDRS-III), in PD patients from Parkinson’s Progression Markers Initiative (PPMI) database ^55^. In fact, the effects of *GCH1* rs11158026 on motor decline of PD patients have been demonstrated by a previous study ^53^. These findings suggested that the motor manifestations of PD patients were significantly shaped by genetic variations. However, the neural mechanisms underlying the effects of these genetic variants on motor function remain largely unknown. We have recently demonstrated that motor and nonmotor symptoms in PD patients were significantly related to brain structural and functional network metrics ^37,55,56^. Additionally, we have shown that the effect of *MAPT* rs17649553 on PD patients’ verbal memory was mediated by the small-worldness metrics of white matter network ^37^. Furthermore, we also reported that the effects of *CCDC62* rs11060180 on motor function of PD patients were mediated by the small-world topology in gray matter covariance network ^55^. Hence, PD risk-associated genetic variants may shape the motor and non-motor symptoms by influencing the structural and functional networks of PD patients ^37,55^.

*BCKDK* rs14235 (G > A) was associated with increased PD risk ^5,6^ and significantly modified the motor function of PD patients ^55^. In this study, we reasoned that *BCKDK* rs14235 may also modify brain network metrics and network metrics shaped by *BCKDK* rs14235 may mediate the effects of *BCKDK* rs14235 on motor function. Therefore, the objective of current study is to evaluate whether *BCKDK* rs14235 affects the structural and functional brain networks of PD patients and to examine whether network metrics shaped by *BCKDK* rs14235 mediate its associations with motor impairment of PD patients. Specifically, our goals consist of: (i) investigate whether *BCKDK* rs14235 A allele is associated scores of motor assessments in PD patients using association analysis; (ii) examine whether structural and functional network measurements vary significantly between GG carriers and A-carriers (GA and AA carriers); (iii) assess whether the effects of *BCKDK* rs14235 on graphical network metrics were independent of common confounding variables, such as age, sex, and disease duration; and (iv) evaluate whether the structural network and functional network measurements mediate the impacts of *BCKDK* rs14235 on motor function of PD patients.

## Materials and Methods

### Participants

The data used in this study were acquired from PPMI cohort study. The PPMI study was approved by Institutional Review Board of each participating site. The informed consents were signed from all participants and could be obtained from the investigators of each participating site. The PD patients were included if they met the criteria below: (i) The patients aged over 30 years old and had a disease duration within 2 years; (ii) The patients had a diagnosis of PD according to the Movement Disorders Society (MDS) Clinical Diagnostic Criteria for PD; (iii) The patients received 3D T1-weighted MPRAGE imaging, resting-state fMRI imaging, and diffusion tensor imaging (DTI) during the same period; (iv) The PD patients received motor assessments including Hoehn and Yahr (H&Y) stages, tremor scores, rigidity scores, and the MDS Unified Parkinson’s Disease Rating Scale part III (MDS-UPDRS). The PD patients were excluded if they met the exclusion criteria below: (i) The patients showed structural abnormalities in the T1-weighted or T2-weighted MRI; (ii) The patients were diagnosed to have PD dementia or dementia with Lewy bodies based on the MDS criteria; (iii) The patients were found to carry the genetic mutations of familial PD or were from the genetic PPMI cohort and prodromal cohort. Finally, one hundred and forty-six PD patients receiving DTI images (n = 146) were included in this study. Among these patients, eighty-three of them (n = 83) also underwent functional MRI. To decipher the associations between *BCKDK* rs14235 (G > A) and motor assessments or brain network metrics, the whole-exome sequencing of blood DNA samples was performed to obtain the genotypes of *BCKDK* rs14235 for individual participant. The demographic and clinical characteristics in different genotype groups (CC carriers, CT carriers, TT carriers) of *BIN3* rs2280104 were compared and shown in Table 1.

**Table 1.** The demographic and clinical data for different genotype groups of *BCKDK* rs14235.

| Clinical variable | GG carriers<br>(n = 46) | A-carriers<br>(n = 100) | GA carriers<br>(n = 83) | AA carriers<br>(n = 17) | GG carriers<br>vs A-carriers | GG carriers vs<br>GA carriers | GG carriers<br>vs AA carriers | Multivariate<br>regression analysis |
| --- | --- | --- | --- | --- | --- | --- | --- | --- |
| Age, years | 62.25 ± 7.75 | 60.78 ± 9.82 | 60.31 ± 10.17 | 63.06 ± 7.74 | $p > 0.05$ | $p > 0.05$ | $p > 0.05$ | - |
| Sex (Male/Female) | 31/15 | 62/38 | 51/32 | 11/6 | $p > 0.05$ | $p > 0.05$ | $p > 0.05$ | - |
| Education, years | 14.91 ± 2.73 | 15.43 ± 3.10 | 15.27 ± 2.97 | 16.24 ± 3.65 | $p > 0.05$ | $p > 0.05$ | $p > 0.05$ | - |
| Disease duration, years | 2.00 ± 1.50 | 2.05 ± 2.40 | 2.08 ± 2.57 | 1.92 ± 1.30 | $p > 0.05$ | $p > 0.05$ | $p > 0.05$ | - |
| HY | 1.68 ± 0.47 | 1.66 ± 0.52 | 1.65 ± 0.53 | 1.71 ± 0.47 | $p > 0.05$ | $p > 0.05$ | $p > 0.05$ | $\beta = -0.0012, p > 0.05$ |
| Tremor | 4.46 ± 3.68 | 3.54 ± 3.03 | 3.51 ± 3.06 | 3.65 ± 2.98 | $p > 0.05$ | $p > 0.05$ | $p > 0.05$ | $\beta = -0.6271, p > 0.05$ |
| Rigidity | 5.32 ± 2.99 | 3.83 ± 2.62 | 3.90 ± 2.65 | 3.47 ± 2.53 | $p < 0.01$ | $p < 0.01$ | $p < 0.05$ | $\beta = -1.0410, p < 0.01$ |
| UPDRS-III | 24.52 ± 10.68 | 19.22 ± 8.71 | 19.29 ± 9.01 | 18.88 ± 7.29 | $p < 0.01$ | $p < 0.01$ | $p < 0.05$ | $\beta = -3.3540, p < 0.01$ |

The data were shown as the mean ± standard deviation (SD). The motor function examination was assessed in ON state. Unpaired t-test (GG carriers vs A-carriers) and one-way ANOVA followed by FDR correction (GG carriers vs GA carrier vs AA carriers) were used to compare the continuous variables among different genotype groups of *BCKDK* rs14235. $\chi^2$ test was used for the comparisons of categorical variables (Sex). Abbreviations: HY, Hoehn & Yahr stage; UPDRS-III, Unified Parkinson' s Disease Rating Scale Part III.

### Image acquisition

The 3D T1-weighted MRI images, resting-state functional images, and DTI images were acquired using 3T Siemens (TIM Trio and Verio) scanners (Erlangen, Germany). The MRI parameters for 3D T1 images were shown below: TR = 2300 ms, TE = 2.98 ms, Voxel size = 1 mm^3^, Slice thickness =1.2 mm, twofold acceleration, sagittal-oblique angulation. The DTI was performed with the parameters as follows: TR= 8,400-8,800 ms, TE = 88 ms, Voxel size = 2 mm^3^, Slice thickness = 2 mm, 64 directions, b = 1000 s/mm^2^. The resting-state functional images were performed using a T2*-weighted echo planar imaging (EPI) sequence as shown below: TR = 2400 ms, TE = 25 ms, Voxel size = 3.3 mm^3^, Slice number = 40, Slice thickness = 3.3 mm, Flip angle = 90°, Acquisition time = 8 min.

## Imaging preprocessing

The DTI images were preprocessed using FMRIB Software Library toolbox (FSL, https://fsl.fmrib.ox.ac.uk/fsl/fslwiki). Briefly, DTI images were corrected for head motions, eddy current distortions, and susceptibility artifacts resulted from magnetic field inhomogeneity. The fractional anisotropy (FA), mean diffusivity (MD), axial diffusivity, and radial diffusivity maps were computed according to the methods as previously reported. The processed images were then reconstructed in standardized MNI space for the following structural network construction.

The SPM12 (http://www.fil.ion.ucl.ac.uk/spm/software/spm12/) and GRETNA software (https://www.nitrc.org/projects/gretna/) were utilized for the preprocessing of resting-state functional images. The detailed preprocessing procedures for functional images included: slice-timing correction, spatial normalizing with standard EPI template, spatial smoothing with 4-mm Gaussian kernel, removal of nuisance signals, and temporal bandpass filtering (0.01–0.1 Hz). The participants (n = 9) with head motion frame-wise displacement (FD) > 0.5 mm and head rotation > 2° were excluded due to excessive head motions according to previous studies ^57–60^. In addition, the head motion parameters were adjusted to correct the effects of head motions on the functional images. Furthermore, we also guaranteed that there were no significant differences in FD values between the GG carriers and T-carriers of *BCKDK* rs14235 (*p* > 0.05).

### Network construction

As previously reported ^37,56^, the white matter network was constructed using deterministic fiber tractography implemented in PANDA (http://www.nitrc.org/projects/panda/), a free and open MATLAB software. The Fiber Assignment by Continuous Tracking (FACT) algorithm was utilized to derive the whole-brain white matter fibers between each pair of 90 nodes in the AAL atlas. The threshold for mean FA skeleton was set to FA > 0.20 and the threshold for white matter fiber angle was set to 45°. As a result, fiber number-based white matter network matrices were constructed in conformity with AAL atlas.

The steps for the construction of functional connectivity matrices were as follows. Firstly, AAL atlas was used to define 90 cortical and subcortical nodes in functional network. Secondly, Pearson correlation coefficient was computed for the time series of 90 nodes to represent the pairwise function connectivity. Finally, the correlation coefficients in functional connectivity matrices were Fisher’s r-to-z transformed to improve the normality of functional connectivity.

### Graph-based network analysis

The GRETNA (https://www.nitrc.org/projects/gretna/) linked with MATLAB software was sued to calculate the graphical network metrics of structural and functional ^61^. During the calculation of global and nodal network metrics, a range of network sparsity thresholds (0.05 ∼ 0.50 with an interval of 0.05) were established. The area under curve (AUC) of global and nodal network metrics was also computed. The global network characteristics consisted of: assortativity, hierarchy, global efficiency, local efficiency, and small-worldness metrics. The small-worldness metrics consisted of: clustering coefficient (Cp), characteristic path length (Lp), normalized clustering coefficient (γ), normalized characteristic path length (λ), and small worldness (σ). The nodal network characteristics include: nodal betweenness centrality, nodal degree centrality, nodal Cp, nodal efficiency, nodal local efficiency, and nodal shortest path length (SPL). The definitions of the aforementioned network metrics have been reported by previous investigations ^55,62,63^.

### Statistical analysis

#### Comparison of clinical variables

For comparisons of continuous variables, unpaired t-test (GG carriers vs A-carriers) and one-way ANOVA test followed by false discovery rate (FDR) correction (GG carriers vs GA carrier vs AA carriers) were used. For comparisons of categorical variables, χ^2^ test was used. *p* < 0.05 was considered statistically significant.

#### Comparison of global network strength

Network-Based Statistic (NBS, https://www.nitrc.org/projects/nbs/) software was used to compare the global network strength of structural and functional networks in different genotype groups of *BCKDK* rs14235 ^64^. The covariates, including age, sex, years of education, and disease duration were included during NBS analysis. *p* < 0.05 after FDR correction was considered statistically significant ^65^.

#### Comparison of network metrics

Two-way ANOVA test followed by FDR corrections was used for the comparisons of global and nodal network metrics among different genotype groups of *BCKDK* rs14235. *p* < 0.05 after FDR correction was considered statistically significant. The AUCs of global network metrics among different genotype groups of *BCKDK* rs14235 were compared using one-way ANOVA test followed by FDR corrections. *p* < 0.05 was considered statistically significant.

#### Association analysis between genotype and scores of motor assessments

The Pearson correlation and multivariate regression analysis were used to analyze the associations between genotype of *BCKDK* rs14235 and scores of motor assessments. During the multivariate regression analysis, age, sex, disease duration, and years of education were included as covariates. *p* < 0.05 was considered statistically significant.

#### Association analysis between genotype and graphical network metrics

The multivariate regression analysis was utilized to analyze the associations between genotype of *BCKDK* rs14235 and graphical network metrics with age, sex, disease duration, and years of education as covariates. FDR-corrected *p* < 0.05 was considered statistically significant.

#### Association analysis between graphical metrics and scores of motor assessments

The Pearson correlation and multivariate regression analysis were used to analyze the associations between graphical metrics and scores of motor assessments. Age, sex, disease duration, and years of education were included as covariates during multivariate regression analysis. *p* < 0.05 was considered statistically significant.

#### Mediation analysis

IBM SPSS Statistics Version 26 was used to perform mediation analysis. The independent variable in the mediation model was the genotype of *BCKDK* rs14235. The dependent variables were the scores of rigidity or UPDRS-III. The mediators were graphical network metrics associated with scores of motor assessments. We modeled the mediated effects of graphical network metrics on the relationships between scores of motor assessments and genotype of *BCKDK*. During the mediation analysis, the age, sex, disease duration, and years of education were included as covariates. *p* < 0.05 was considered statistically significant.

## Results

### Allele and genotype frequencies

The allele frequencies (G allele frequency = 0.5993, A allele frequency = 0.4007) for *BCKDK* rs14235 were comparable to the data from Allele Frequency Aggregator Project (A allele frequency = 0.3798). The distribution of genotype frequencies from our patients didn’t deviate Hardy–Weinberg equilibrium (*p* > 0.10). Genotype frequencies were similar with respect to age or sex (*p* > 0.05).

### Group difference of clinical variables and multivariate regression analysis

The group differences of clinical variables in different genotype groups of *BCKDK* rs14235 were shown in Table 1 and Figure 1. The Hoehn & Yahr stages and tremor scores were not statistically different between GG carriers and A-carriers (GA carriers and AA carriers). The rigidity scores and UPDRS-III scores were significantly lower in A-carriers compared to GG carriers (Table 1 and Figure 1). Consistently, the rigidity scores and UPDRS-III scores in GA carriers and AA carriers were also much lower than those of GG carriers (Table 1 and Figure 1). The *BCKDK* rs14235 A allele was negatively associated with rigidity scores (β = −1.0410, *p* < 0.01) and UPDRS-III scores (β = −3.3540, *p* < 0.01).

**Figure 1.**
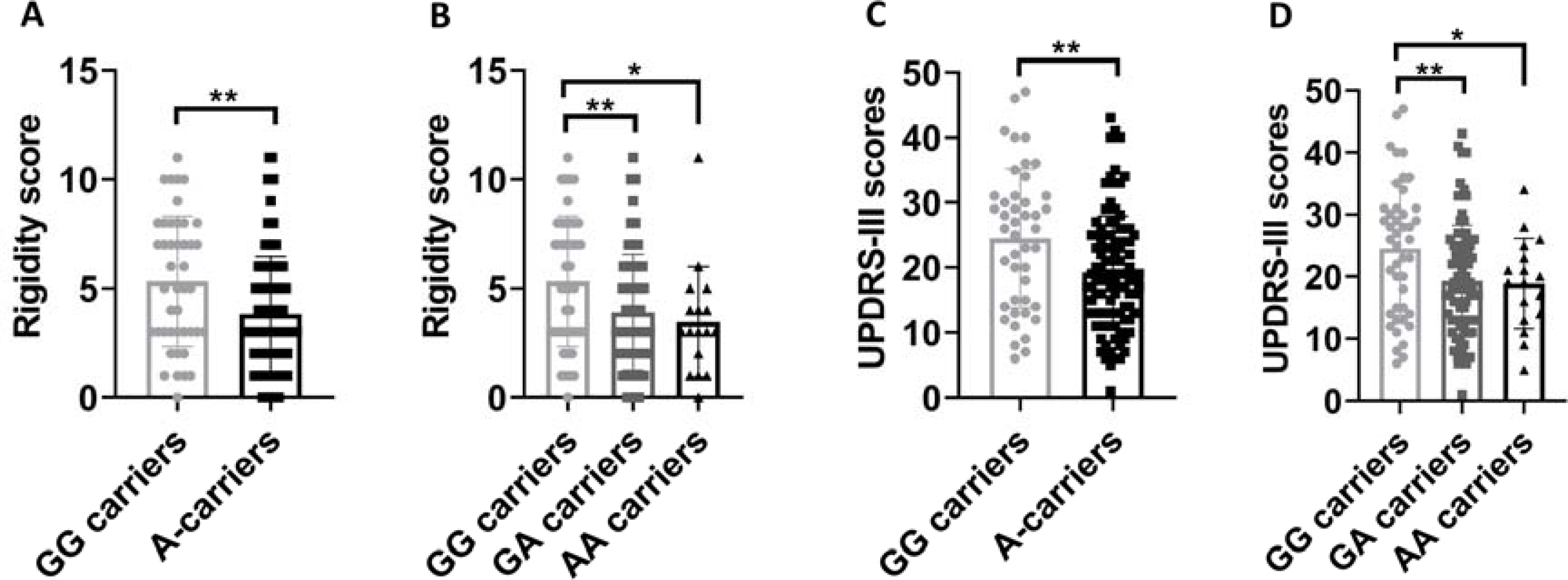
Group differences of motor assessments between GG carriers and A-carriers of PD patients. (A) A-carriers exhibited lower rigidity scores compared to GG carriers. (B) GA and AA carriers showed lower rigidity scores compared to GG carriers. (C) A-carriers presented lower UPDRS-III scores compared to GG carriers. (D) GA and AA carriers showed lower UPDRS-III scores compared to GG carriers. Unpaired t-test (GG carriers *vs* A-carriers) and one-way ANOVA test followed by FDR corrections (GG carriers *vs* GA carrier *vs* AA carriers) were used. *p* < 0.05 was considered statistically significant. Abbreviations: UPDRS-III, Unified Parkinson’s Disease Rating Scale part III.

### Group difference of graphical network metrics in structural network

Compared to GG carriers and GA carriers, AA carriers exhibited higher assortativity (FDR-corrected *p* < 0.05, sparsity range: 0.10 – 0.50; Figure 2A). Compared to GG carriers, AA carriers had lower small-worldness γ and σ (FDR-corrected *p* < 0.05, sparsity range: 0.15,0.30, 0.45-0.50; Figure 2A-C). Consistently, we also found higher AUC of assortativity in AA carriers compared to GG carriers and GA carriers (FDR-corrected *p* < 0.05; Figure S1A). Additionally, AA carriers exhibited lower AUCs of small-worldness γ and σ compared to GG and GA carriers (FDR-corrected *p* < 0.05; Figure S1B-C). For nodal metrics, AA carriers exhibited lower betweenness centrality in bilateral calcarine, left precuneus, and left thalamus compared to GG and GA carriers (FDR-corrected *p* < 0.05; Figure 2D). Compared to GG carriers, GA and AA carriers showed higher betweenness centrality, degree centrality, and nodal efficiency in right precuneus (FDR-corrected *p* < 0.05; Figure 2D, E, G). In addition, some key nodes in basal ganglia network, such as left putamen, right putamen, right pallidum also showed differential nodal network metrics among different genotype groups of *BCKDK* rs14235 (FDR-corrected *p* < 0.05; Figure 2D, E, G, I). Interestingly, AA carriers showed higher nodal Cp, nodal efficiency, and nodal local efficiency in right heschl compared to GG and GA carriers (FDR-corrected *p* < 0.05; Figure 2F-H).

**Figure 2.**
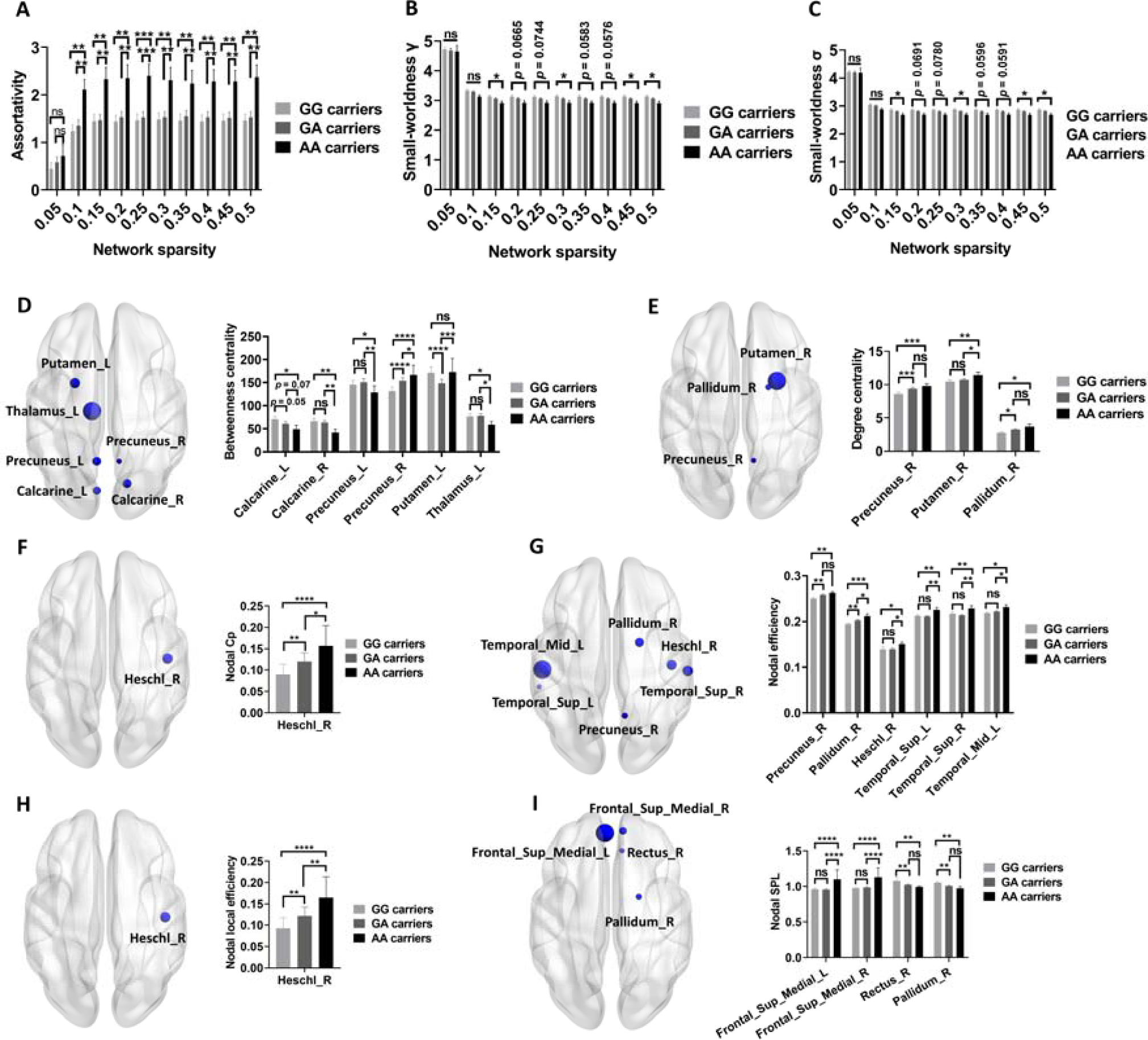
Group differences of structural network metrics among GG, GA, and AA carriers. (A-C) Group differences in assortativity (A), small-worldness _γ_ (B), and small-worldness _σ_ (C) among GG, GA, and AA carriers (FDR-corrected *p* < 0.05, Two-way ANOVA test). (D-I) Group differences in nodal betweenness centrality (D), degree centrality (E), Cp (F), efficiency (G), local efficiency (H), and SPL (I) among GG, GA, and AA carriers (FDR-corrected *p* < 0.05, Two-way ANOVA test). Two-way ANOVA test followed by FDR corrections (GG carriers *vs* GA carrier *vs* AA carriers) were used. *p* < 0.05 was considered statistically significant. Abbreviations: Cp, Clustering coefficient; _γ_, normalized clustering coefficient; _σ_, small worldness; SPL, shortest path length.

### Group difference of graphical network metrics in functional network

Compared to GG carriers, GA and AA carriers exhibited higher global efficiency (FDR-corrected *p* < 0.05, sparsity range: 0.10-0.20; Figure 3A) and local efficiency (FDR-corrected *p* < 0.05, sparsity range: 0.05-0.15; Figure 3B) at multiple sparsity thresholds. In agreement with this, the AUCs of global efficiency and local efficiency were also higher in GA and AA carriers compared to GG carriers (FDR-corrected *p* < 0.05; Figure S1D-E). For nodal network metrics, AA carriers showed higher betweenness centrality in right middle fontal gyrus, right posterior cingulate gyrus, and bilateral precuneus compared to GA and GG carriers (FDR-corrected *p* < 0.05; Figure 3C). Moreover, AA carriers had higher degree centrality in right superior orbitofrontal gyrus and right posterior cingulate gyrus compared to GG and GA carriers (FDR-corrected *p* < 0.05; Figure 3D). Interestingly, network metrics of key nodes in basal ganglia network, including right caudate, left caudate, and right putamen were specifically modified by *BCKDK* rs14235 (FDR-corrected *p* < 0.05; Figure 3E-H). Furthermore, AA carriers exhibited higher nodal efficiency in bilateral posterior cingulate gyrus compared to GG carriers, companied by lower nodal SPL in AA carriers compared to GG carriers (FDR-corrected *p* < 0.05; Figure 3E and H). Taken together, these data supported that *BCKDK* rs14235 A allele enhanced the nodal network metrics of functional networks in PD.

**Figure 3.**
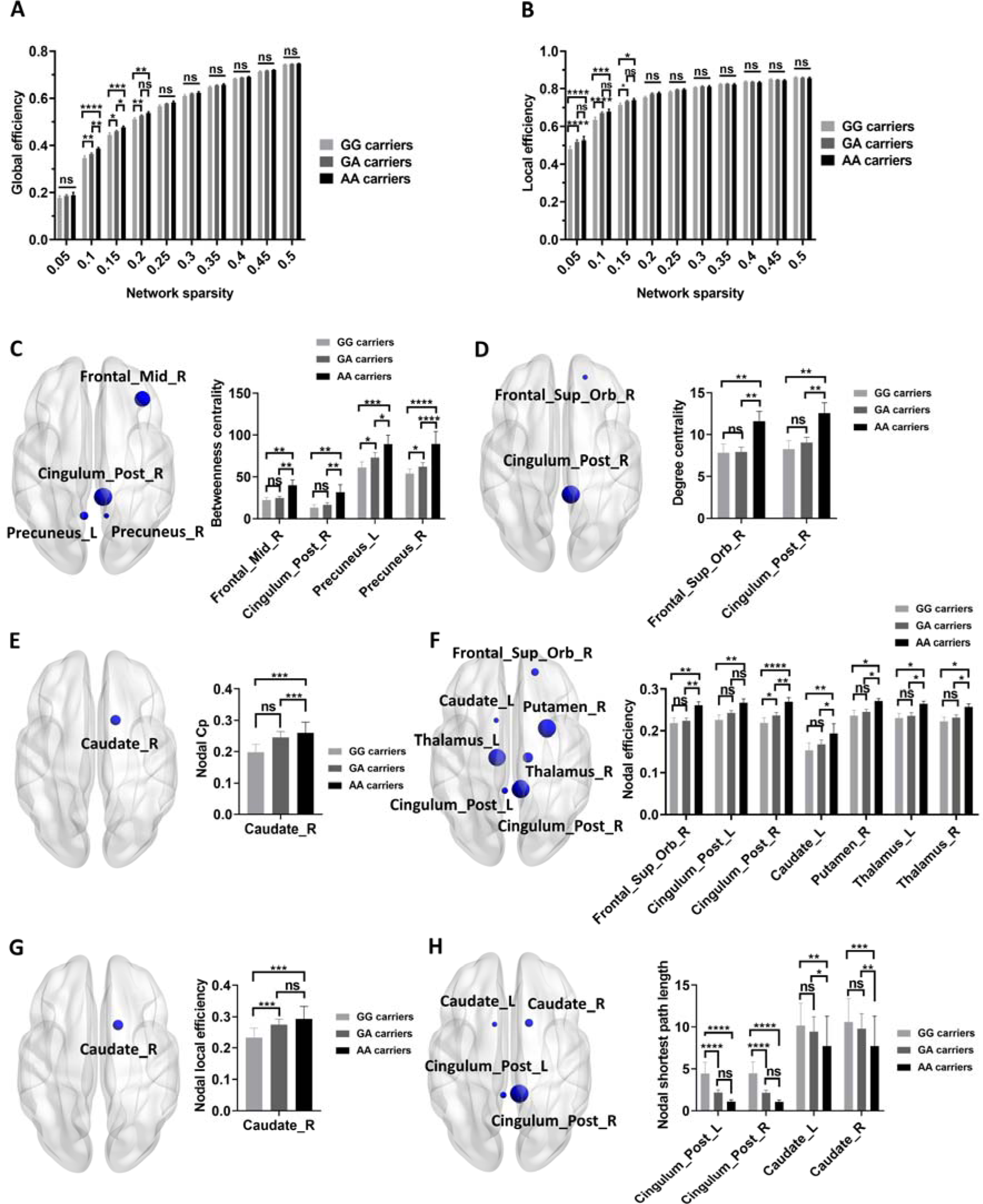
Group differences of functional network metrics among GG, GA, and AA carriers. (A-B) Group differences in global efficiency (A) and local efficiency (B) among GG, GA, and AA carriers (FDR-corrected *p* < 0.05, Two-way ANOVA test). (D-I) Group differences in nodal betweenness centrality (C), degree centrality (D), Cp (E), efficiency (F), local efficiency (G), and SPL (H) among GG, GA, and AA carriers (FDR-corrected *p* < 0.05, Two-way ANOVA test). Two-way ANOVA test followed by FDR corrections (GG carriers *vs* GA carrier *vs* AA carriers) were used. *p* < 0.05 was considered statistically significant. Abbreviations: Cp, Clustering coefficient; SPL, shortest path length.

### The associations between *BCKDK* rs14235 and graphical network metrics

To examine whether the effects of *BCKDK* rs14235 on graphical network metrics were independent of common confounding variables, multivariate regression analysis was used to assess the associations between *BCKDK* rs14235 and graphical network metrics showing statistical group difference in Figure 2-3. As shown in Table S1 and Table S2, *BCKDK* rs14235 was significantly associated with multiple brain network metrics in both structural and functional networks (FDR-corrected *p* < 0.05; Table S1 and Table S2). These results demonstrated that *BCKDK* rs14235 independently shaped the network topology of both structural and functional network.

### Mediation analysis

To explore whether graphical network metrics contributed to the effects of *BCKDK* rs14235 on motor impairment of PD patients, we analyzed the associations between graphical network metrics and rigidity scores or UPDRS-III scorers. Among those metrics showing significant group differences, AUC of assortativity in structural network was negatively associated with scores of rigidity (β = −1.29, *p* < 0.05; Figure 4A) while nodal SPL of right pallidum in structural network was positively associated with scores of UPDRS-III (β = 29.23, *p* < 0.01; Figure 4B). With mediation analysis, we further demonstrated that increased AUC of assortativity in structural network mediated the effects of *BCKDK* rs14235 on rigidity (Figure 4C). Additionally, lower nodal SPL in right pallidum in structural network mediated the effects of *BCKDK* rs14235 on UPDRS-III scores (Figure 4D).

**Figure 4.**
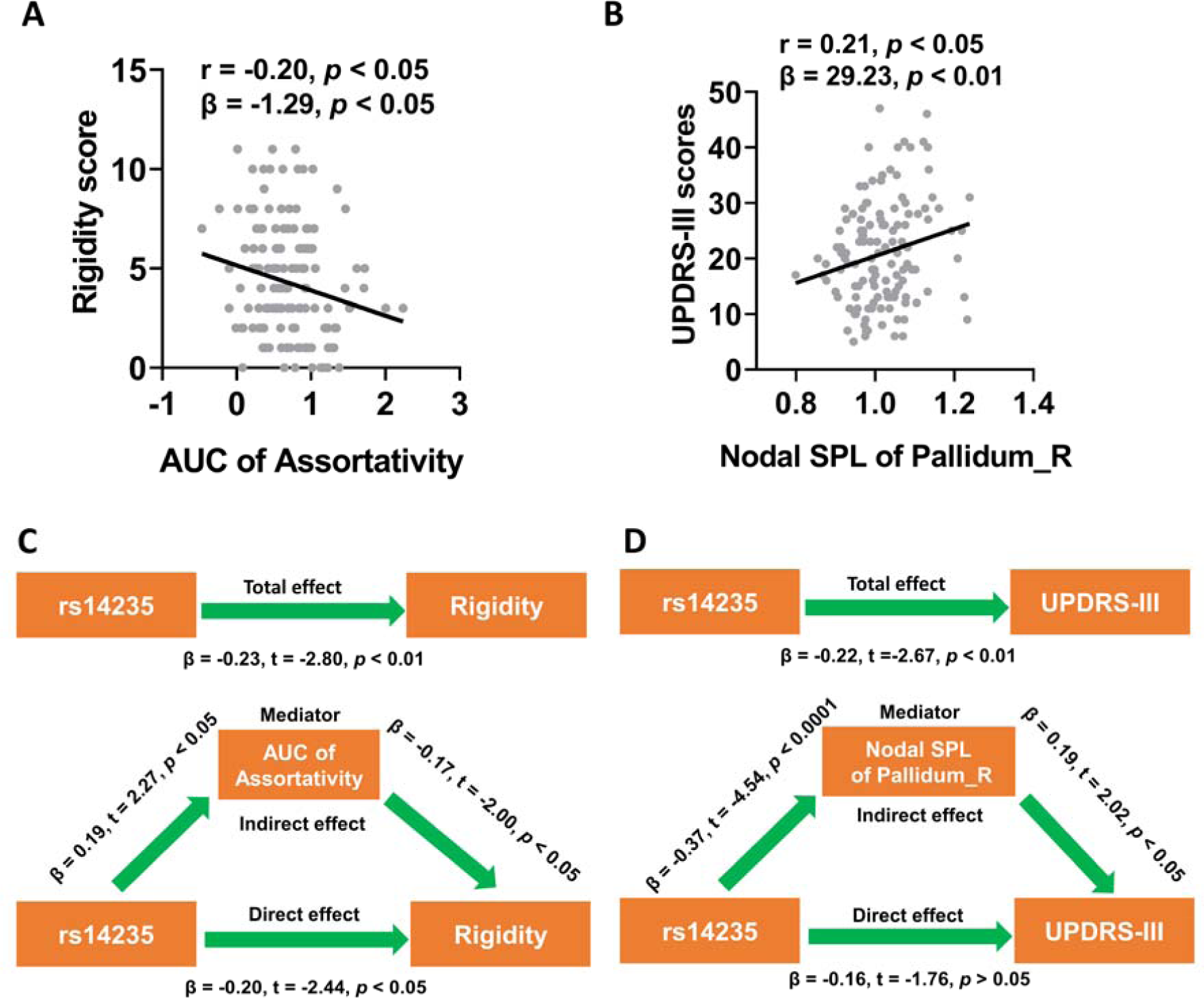
Mediation analysis suggested structural network metrics mediated the effects of *BCKDK* rs14235 on motor assessments of PD patients. (A) AUC of assortativity was negatively associated with the rigidity scores of PD patients (*p* < 0.05 in both Pearson correlation analysis and multivariate regression analysis). (B) Nodal SPL of right pallidum was positively associated with UPDRS-III scores in PD patients (*p* < 0.05 in both Pearson correlation analysis and multivariate regression analysis). (C) AUC of assortativity mediated the effects of *BCKDK* rs14235 on rigidity scores. (D) Nodal SPL of right pallidum mediated the effects of *BCKDK* rs14235 on UPDRS-III scores. The association analysis between network metrics and scores of motor assessments was conducted by Pearson correlation method and multivariate regression analysis with age, sex, disease duration, and years of education as covariates. During the mediation analysis, age, sex, disease duration, and years of education were included as covariates. *p* < 0.05 was considered statistically significant. Abbreviations: AUC, Area under curve; UPDRS-III, Unified Parkinson’s Disease Rating Scale part III; SPL, shortest path length.

## Discussion

In this study, we revealed that a PD-associated risk variant, *BCKDK* rs14235, was outstandingly associated with scores of rigidity and UPDRS-III. In addition, *BCKDK* rs14235 prominently affected the network metrics of structural and functional network. Furthermore, differential structural network metrics were correlated with rigidity scores and UPDRS-III scores, respectively. Finally, we demonstrated that it was the increased AUC of assortativity and lower nodal SPL in right pallidum in structural network respectively mediated the effects of *BCKDK* rs14235 on rigidity scores and UPDRS-III scores.

### The effects of *BCKDK* rs14235 on motor decline of PD patients

There is increasing consensus that PD is a neurodegenerative disorder with heterogeneous motor and non-motor features ^28–31^. According to previous literature, the heterogeneity of clinical manifestations was associated with age ^56,66^, sex ^56,66^, life styles ^28,67,68^, genetic variations ^32,38,44,69^, β-amyloid deposition ^70^, tau accumulation ^70,71^, varied pathological spread of synucleinopathy ^28,70^ and diverse degenerative thresholds ^28,72^. Genetic variations play an essential role in the modifications of clinical phenotypes of PD patients. Particularly, numerous mutated genes in PD, such as *GBA* ^36,38,39^, *LRRK2* ^24,40,41^, *PRKN* ^42,43^, *PINK1* ^43,44^, *SNCA* ^41,45^ have been demonstrated to significantly shape the clinical heterogeneity of familial PD patients. Motor symptoms of PD patients, on the other hand, were also influenced by genetic variations. It has been previously shown that the motor impairment of PD patients were linked to a series of genetic variants, including *GBA* ^46,47^, *SNCA* ^48,49^, *BST1* ^50^, *ATP8B2* ^51^, *PARK16* ^52^, *GCH1* ^53^, and *APOE* ^54^. Recently, we identified 3 genetic variants, *CCDC62* rs11060180, *GCH1* rs11158026 and *BCKDK* rs14235, were significantly associated with UPDRS-III scores in 198 PD patients ^55^. In current study, we confirmed that *BCKDK* rs14235 A allele was negatively associated with UPDRS-III scores in 146 PD patients. In addition, *BCKDK* rs14235 A allele was also associated with milder rigidity in PD patients, which further supported that *BCKDK* rs14235 specifically modified motor function of PD patients ^55^. Interestingly, *BCKDK* rs14235 A allele was not associated with tremor scores, suggesting that the effect of *BCKDK* rs14235 A allele was only limited to parts of motor symptoms, especially rigidity. Previous studies have reported that *GCH1* rs11158026 T allele was associated with the impairment of dopaminergic innervation in striatum, which might help to explain why this allele was associated with higher UPDRS-III scores ^53,55^. In contrast, *BCKDK* rs14235 was not found to be associated with striatal binding ratios (SBRs) of bilateral striatum of PD patients ^55^, suggesting that the effects of *BCKDK* rs14235 on motor assessments were independent of striatum SBRs. Taken together, we concluded that *BCKDK* rs14235 was a PD risk-associated genetic variant specifically shaping the motor impairment of PD patients.

### The effects of *BCKDK* rs14235 on structural network metrics

We found *BCKDK* rs14235 significantly modified the structural network metrics of PD patients. Compared to GG and GA carriers, AA carriers showed higher assortativity. Assortativity represents the tendency of nodes to connect with ‘similar’ nodes over ‘dissimilar’ nodes in a network. It has been shown that assortativity determined the robustness of a network to noise. For example, robustness is greatly enhanced in assortative neural networks ^73^. Additionally, robustness exhibits a noticeable increase as assortativity becomes more positive in gene regulatory networks ^74^. Furthermore, assortativity was also associated with higher synchronization in complex neuronal network ^75^.In current study, we found AA carriers with milder motor decline exhibited higher assortativity compared to GG and GA carriers, which suggested that AA carriers might have higher robustness to noise or random attacks due to their higher network assortativity. However, according to previous studies, PD patients seemed to exhibit increased assortativity ^76–78^, indicating higher assortativity may be associated with worse motor impairment in PD. One explanation for this discrepancy may be that these studies investigated the functional network and structural covariance network but not white matter network, which usually exhibited distinct topological properties compared to functional and structural covariance network ^79–82^. Small-world properties have been discovered in a multitude of biological networks, including gene regulatory network ^83^, microRNA functional networks ^84^, protein interaction network ^85^, and neural networks ^86^. Recently, we found *MAPT* rs17649553 T allele was associated with higher small-worldness properties in structural network and enhanced small-world topology in structural network mediated the effects of *MAPT* rs17649553 T allele on verbal memory of PD patients ^37^. In this study, we found AA carriers exhibited lower small-worldness γ and σ compared to GG carriers, indicating that small-world topology of structural network was specifically targeted by PD-associated risk variants ^55^. For nodal network metrics, we found network metrics of some key nodes in default mode network and basal ganglia network were preferentially modified. Precuneus was a key node in default mode network and was revealed to be impaired in PD patients. For example, precuneus atrophy and hypometabolism has been found in PD patients with isolated apathy ^87^. The disruption of functional connectivity and blood perfusion in precuneus was also reported by a series of studies ^88–90^. We found higher betweenness centrality, degree centrality, and nodal efficiency in right precuneus of AA carriers compared to GG carriers, indicating that local topology in right precuneus was specifically modulated by A allele of *BCKDK* rs14235. We found nodal network metrics in multiple key nodes of basal ganglia network, such as left putamen, right putamen, and right pallidum, were significantly modified, implying that basal ganglia network was preferentially regulated by A allele of *BCKDK* rs14235. Indeed, the alterations of nodal network metrics in putamen and pallidum have been revealed in PD patients ^91,92^. For instance, remarkable decreases of nodal efficiency have been observed in globus pallidus and putamen of PD patients ^92^. Recently, we showed that *MAPT* rs17649553 and *BIN3* rs2280104 significantly modified betweenness centrality of left putamen ^37,93^, which indicated that left putamen was a shared target of PD-associated risk variants. Taken together, our findings demonstrated that structural network metrics were prominently shaped by *BCKDK* rs14235.

### The effects of *BCKDK* rs14235 on functional network metrics

In addition to structural network, functional network was also significantly affected by *BCKDK* rs14235. We found higher global efficiency and local efficiency in AA and GA carriers compared to GG carriers, which suggested *BCKDK* rs14235 specifically modified network efficiency of functional network. In fact, compared to control participants, network local efficiency was significantly reduced in PD patients ^94,95^, whereas network global efficiency seemed to be increased in PD patients ^95,96^. Therefore, our findings indicated that *BCKDK* rs14235 might contribute to the disturbances of network efficiency in PD patients. We found nodal metrics in caudate, putamen, precuneus, and posterior cingulate gyrus were significantly modified by *BCKDK* rs14235, which suggested that similar to structural network, *BCKDK* rs14235 also preferentially shaped the local topology of basal ganglia network and default mode network in functional network of PD patients. The alterations of basal ganglia network and default mode network in PD have been demonstrated by several investigations ^92,97–100^. Thus, *BCKDK* rs14235 specifically shaped PD-related networks, especially basal ganglia network and default mode network. Interestingly, AA carriers showed consistently higher nodal betweenness centrality, degree centrality, nodal Cp, nodal efficiency, nodal local efficiency, and lower nodal SPL compared to GG and GA carriers, which suggested *BCKDK* rs14235 A allele was associated with enhanced functional network topology in comparison with G allele. In a recent study, we found *BIN3* rs2280104 T-carriers exhibited lower nodal Cp, efficiency, local efficiency, and higher shortest path length in left caudate ^93^. In this study, we found higher nodal Cp, nodal local efficiency and lower nodal SPL in right caudate of AA carriers, which suggested local topology of caudate was specifically shaped by PD-associated risk variants^93^.Taken together, our findings demonstrated that functional network metrics were also dramatically modified by *BCKDK* rs14235.

### The associations between network metrics and scores of motor assessments

We found structural network metrics were significantly associated with motor assessments of PD patients, indicating that structural network metrics were potential predictors of motor function in PD. In a recent study, we reported 3 genetic variants, *GCH1* rs11158026, *CCDC62* rs11060180, and *BCKDK* rs14235, were significantly correlated with UPDRS-III scores in PD patients ^55^, nevertheless, the neural mechanisms underlying the effects of these variants on UPDRS-III scores were not identified. According to a previous study, it seemed that *GCH1* rs11158026 T allele contributed to the deterioration of motor function via the disruption of dopaminergic innervation in striatum ^53^. For *CCDC62* rs11060180, we have recently demonstrated that small-world topology in gray matter covariance network mediated the effects of *CCDC62* rs11060180 on motor assessments of PD patients ^55^. In this study, we showed that the lower nodal SPL of right pallidum, a key node in basal ganglia network, mediated the effects of *BCKDK* rs14235 on UPDRS-III scores. Pallidum, namely globus pallidus, was a core node in basal ganglia network, which played a key role in the occurrence of PD ^101,102^. In addition, pallidum was also an important target of neuromodulation in PD ^103,104^. Furthermore, the reduction of nodal efficiency in pallidum has been reported in PD patients, indicating impaired local information transfer efficiency in PD ^92^. In this study, we found significantly differential nodal network metrics in right pallidum among GG carriers, GA carriers, and AA carriers, characterized by higher nodal degree centrality and nodal efficiency, as well as lower nodal SPL in A-carriers. These results suggested that A-carriers exhibited higher information transfer efficiency in right pallidum compared to GG carriers, which provided reliable explanations at the network level for the negative association between *BCKDK* rs14235 A allele and UPDRS-III scores in PD patients. To summarize, we concluded that *BCKDK* rs14235 A allele delayed motor decline in PD by improving the local information transfer of right pallidum of structural network.

### The potential molecular mechanisms underlying the effects of *BCKDK* rs14235

Branched-chain ketoacid dehydrogenase kinase (BCKDK) is a key rate-limiting enzyme catalyzing the oxidation of branched-chain amino acids ^105,106^. BCKDK was associated with a multitude of diseases, including cancer ^105–107^, nonalcoholic fatty liver disease ^108^, neurodevelopmental diseases ^109–112^, heart failure ^113,114^, and also PD ^115,116^. In this study, we found a genetic variant of *BCKDK* gene, rs14235 was significantly associated with motor assessments and network topology of PD patients, which suggested that BCKDK or branched-chain amino acid metabolism may be involved in the pathogenesis of PD. According to GTEx database, *BCKDK* rs14235 was associated with differential expressions of multiple genes in brain tissue, including *KAT8*, *ZNF646*, *VKORC1*, and *BCKDK* ^55^. Dang et al. (2022) revealed that *KAT8* gene was causally associated with PD using dopaminergic neuron-specific eQTL in a mendelian randomization study ^117^. KAT8 has been shown to regulate PINK1-dependent mitophagy initiation and partially underlined the genetic risk for PD at 16q11.2 and *MAPT* H1 loci ^118^. Zinc finger (ZNF) gene family has been recently demonstrated to be associated with PD ^119^, such as *ZNF219* and *ZNF746*, however, whether *ZNF646* conferred a risk of PD remained elusive. *VKORC1* gene has been shown to be associated with ischemic stroke ^120^, vascular dementia ^121^, and Alzheimer’s disease ^122^, whereas whether it was also associated with PD was unknown. On the basis of these findings, we concluded that the role of *BCKDK* gene in PD pathogenesis remained largely unknown. Future studies were required to decipher how *BCKDK* rs14235 affected disease risk, motor function, and network topology of PD patients.

### Strengths and limitations of this study

In this study, we reported a PD-associated risk variant, *BCKDK* rs14235 was significantly associated with motor assessments and network topology of PD patients. This finding provided potential interpretations for the individual heterogeneity of motor symptoms in PD patients. In addition, our results also suggested that *BCKDK* rs14235 A allele is a genetic predictor of motor function in PD patients. The effects of *BCKDK* rs14235 on structural network and functional network indicated that *BCKDK* rs14235 significantly shaped the topological organizations of brain network. Particularly, *BCKDK* rs14235 A allele enhanced the local information transfer of right pallidum of structural network, which mediated the effects of *BCKDK* rs14235 A allele on motor assessment of PD patients. This provided plausible network mechanisms to explain the effects of *BCKDK* rs14235 A allele on motor decline of PD patients. The limitation of this study was that we did not determine the molecular mechanisms mediating the effects of *BCKDK* rs14235 on motor function and brain networks. Further studies are encouraged to decipher the impact of *BCKDK* rs14235 on PD pathogenesis at the molecular level.

## Conclusions

To summarize, we reported a PD risk-associated genetic variant, *BCKDK* rs14235, was significantly associated with motor assessments of PD patients. In addition, we found *BCKDK* rs14235 A allele modified both structural network and functional network. Moreover, we further showed structural network metrics were significantly correlated with motor symptoms. Finally, we demonstrated that it was the local topology of right pallidum that mediated the effects of *BCKDK* rs14235 A allele on motor assessments of PD patients.

## Author contributions

Zhichun Chen, Conceptualization, Formal analysis, Visualization, Methodology, Writing - original draft, Writing - review and editing; Bin Wu, Validation, Investigation, Methodology; Guanglu Li, Data curation, Formal analysis, Visualization; Liche Zhou, Data curation, Formal analysis, Investigation; Lina Zhang, Formal analysis, Investigation, Methodology; Jun Liu, Conceptualization, Supervision, Funding acquisition, Writing - original draft, Project administration, Writing-review and editing.

## Acknowledgments

Data used in the preparation of this article were obtained from the Parkinson’s Progression Markers Initiative (PPMI) database (www.ppmiinfo.org/data). We thank the share of PPMI data by all the PPMI study investigators. PPMI – a public-private partnership – is funded by the Michael J. Fox Foundation for Parkinson’s Research and funding partners, which can be found at www.ppmiinfo.org/fundingpartners.

## Data availability

All the raw data used in the preparation of this Article were downloaded from PPMI database (www.ppmi-info.org/data). All data produced in the present study are available upon reasonable request to the authors.

## Supporting Information

Additional supporting information may be found online in the Supporting Information section at the end of the article.

