## Supplemental file for "*BCKDK* rs14235 A allele is associated with milder motor impairment and altered network topology in Parkinson’s disease"

**Table S1** The associations between *BCKDK* rs14235 and graphical metrics of structural network

| Assortativity | | | | | | | | | | | | | | | | | | | | | | | | | | | | | | | | | |
| --- | --- | --- | --- | --- | --- | --- | --- | --- | --- | --- | --- | --- | --- | --- | --- | --- | --- | --- | --- | --- | --- | --- | --- | --- | --- | --- | --- | --- | --- | --- | --- | --- | --- |
| Sparsity=0.1 | | | Sparsity=0.15 | | | | Sparsity=0.2 | | | | Sparsity=0.25 | | | | Sparsity=0.3 | | | | Sparsity=0.35 | | | | Sparsity=0.4 | | | | Sparsity=0.45 | | | | Sparsity=0.5 | | |
| β = 0.33  *p* = 0.0941 | | | β = 0.30  **p* = 0.0473 | | | | β = 0.33  *p =* 0.0675 | | | | β = 0.33  *p* = 0.1746 | | | | β = 0.29  *p* = 0.0523 | | | | β = 0.28  **p* = 0.0476 | | | | β = 0.31  *p* = 0.05652 | | | | β = 0.29  **p* = 0.0461 | | | | β = 0.32  *p* = 0.0569 | | |
| **Small-worldness γ** | | | | | | | | | | | | | | | | | | | | | | | | | | | | | | | | | |
| Sparsity=0.15 | | | | Sparsity=0.2 | | | | Sparsity=0.25 | | | | | Sparsity=0.3 | | | | Sparsity=0.35 | | | | Sparsity=0.4 | | | | | Sparsity=0.45 | | | | Sparsity=0.5 | | | |
| β = -0.11  **p* = 0.0131 | | | | β = -0.10  ***p* = 0.0099 | | | | β = -0.10  ***p =* 0.0096 | | | | | β = -0.11  **p* = 0.0102 | | | | β = -0.10  ***p* = 0.0085 | | | | β = -0.10  **p* = 0.0101 | | | | | β = -0.11  **p* = 0.0344 | | | | β = -0.11  **p =* 0.0184 | | | |
| **Small-worldness σ** | | | | | | | | | | | | | | | | | | | | | | | | | | | | | | | | | |
| Sparsity=0.15 | | | | Sparsity=0.2 | | | | Sparsity=0.25 | | | | | Sparsity=0.3 | | | | Sparsity=0.35 | | | | Sparsity=0.4 | | | | | Sparsity=0.45 | | | | Sparsity=0.5 | | | |
| β = -0.09  **p* = 0.0131 | | | | β = -0.08  **p* = 0.0106 | | | | β = -0.08  **p =* 0.0102 | | | | | β = -0.09  **p* = 0.0102 | | | | β = -0.08  ***p* = 0.0087 | | | | β = -0.08  **p* = 0.0102 | | | | | β = -0.09  **p* = 0.0344 | | | | β = -0.09  **p =* 0.0184 | | | |
| **Nodal betweenness centrality** | | | | | | | | | | | | | | | | | | | | **Nodal degree centrality** | | | | | | | | | | | | | **Nodal Cp** |
| Calcarine_L | | Calcarine_R | | | | Precuneus_L | | | | Precuneus_R | | | | Putamen_L | | | Thalamus_L | | | Precuneus_R | | | | Putamen_R | | | | Pallidum_R | | | | | Heschl_R |
| β = -10.39  *p* = 0.1224 | | β = -8.94  *p* = 0.1368 | | | | β = -2.87  *p* = 0.7443 | | | | β = 16.91  *p* = 0.2358 | | | | β = -5.25  *p* = 0.7496 | | | β = -5.95  *p* = 0.4578 | | | β = 0.61  **p* = 0.0156 | | | | β = 0.46  **p* = 0.0295 | | | | β = 0.53  ****p* = 0.0009 | | | | | β = 0.03  *p* = 0.1708 |
| **Nodal efficiency** | | | | | | | | | | | | | | | | | | **Nodal local efficiency** | | | | **Nodal shortest path length** | | | | | | | | | | | |
| Precuneus_R | Pallidum_R | | | | Heschl_R | | | | Temporal_Sup_L | | | Temporal_Sup_R | | | | Temporal_Mid_L | | Heschl_R | | | | Frontal_Sup_Medial_L | | | Frontal_Sup_Medial_R | | | | Rectus_R | | | Pallidum_R | |
| β = 0.0068  ***p* = 0.009 | β = 0.0093  ****p* = 0.0001 | | | | β = 0.0052  *p* = 0.17976 | | | | β = 0.0045  *p* = 0.0908 | | | β = 0.0037  *p* = 0.1865 | | | | β = 0.0063  ***p* = 0.0072 | | β = 0.0343  *p* = 0.1629 | | | | β = 0.0442  *p* = 0.1033 | | | β = 0.0547  *p* = 0.1104 | | | | β = -0.0486  *p* = 0.1175 | | | β = -0.0466  *****p* < 0.0001 | |

Multivariate regression analysis was performed by adjusting age, sex, disease duration, and years of education. False discovery rate (FDR) was used for multiple comparison corrections. FDR-corrected *p*-values were shown. FDR-corrected *p* < 0.05 was considered statistically significant. **p* < 0.05, ***p* < 0.01, ****p* < 0.001, *****p* < 0.0001.

**Table S2** The associations between *BCKDK* rs14235 and graphical metrics of functional network

| Global efficiency | | | | | | | | Local efficiency | | | | | | | |
| --- | --- | --- | --- | --- | --- | --- | --- | --- | --- | --- | --- | --- | --- | --- | --- |
| Sparsity=0.1 | | Sparsity=0.15 | | | Sparsity=0.2 | | | Sparsity=0.05 | | | Sparsity=0.10 | | | Sparsity=0.15 | |
| β = 0.02  ***p* = 0.0071 | | β = 0.02  ***p* = 0.0057 | | | β = 0.01  **p* = 0.0105 | | | β = 0.03  **p* = 0.0312 | | | β = 0.02  **p* = 0.0354 | | | β = 0.02  **p* = 0.0237 | |
| **Nodal betweenness centrality** | | | | | | | | | **Nodal degree centrality** | | | | | | **Nodal Cp** |
| Frontal_Mid_R | Cingulum_Post_R | | | Precuneus_L | | Precuneus_R | | | Frontal_Sup_Orb_R | | | Cingulum_Post_R | | | Caudate_R |
| β = 7.32  **p* = 0.0464 | β = 8.20  **p* = 0.0201 | | | β = 12.74  *p* = 0.0520 | | β = 15.58  **p* = 0.0248 | | | β = 1.69  **p* = 0.0438 | | | β = 1.84  **p* = 0.0299 | | | β = 0.04  *p* = 0.0897 |
| **Nodal efficiency** | | | | | | | | | | | | | | | |
| Frontal_Sup_Orb_R | Cingulum_Post_L | | | Cingulum_Post_R | | Caudate_L | | | Putamen_R | | | Thalamus_L | | | Thalamus_R |
| β = 0.02  **p* = 0.0436 | β = 0.02  **p* = 0.0473 | | | β = 0.02  **p* = 0.0462 | | β = 0.02  *p* = 0.1750 | | | β = 0.01  *p* = 0.1417 | | | β = 0.01  *p* = 0.1610 | | | β = 0.01  *p* = 0.1395 |
| **Nodal local efficiency** | | | **Nodal shortest path length** | | | | | | | | | | | | |
| Caudate_R | | | Cingulum_Post_L | | | | Cingulum_Post_R | | | Caudate_L | | | Caudate_R | | |
| β = 0.03  *p* = 0.2223 | | | β = -1.85  **p* = 0.0315 | | | | β = -1.88  *p* = 0.0560 | | | β = -1.21  *p* = 0.7033 | | | β = -1.20  *p* = 0.5726 | | |

Multivariate regression analysis was performed by adjusting age, sex, disease duration, and years of education. False discovery rate (FDR) was used for multiple comparison corrections. FDR-corrected *p*-values were shown. FDR-corrected *p* < 0.05 was considered statistically significant. **p* < 0.05, ***p* < 0.01.

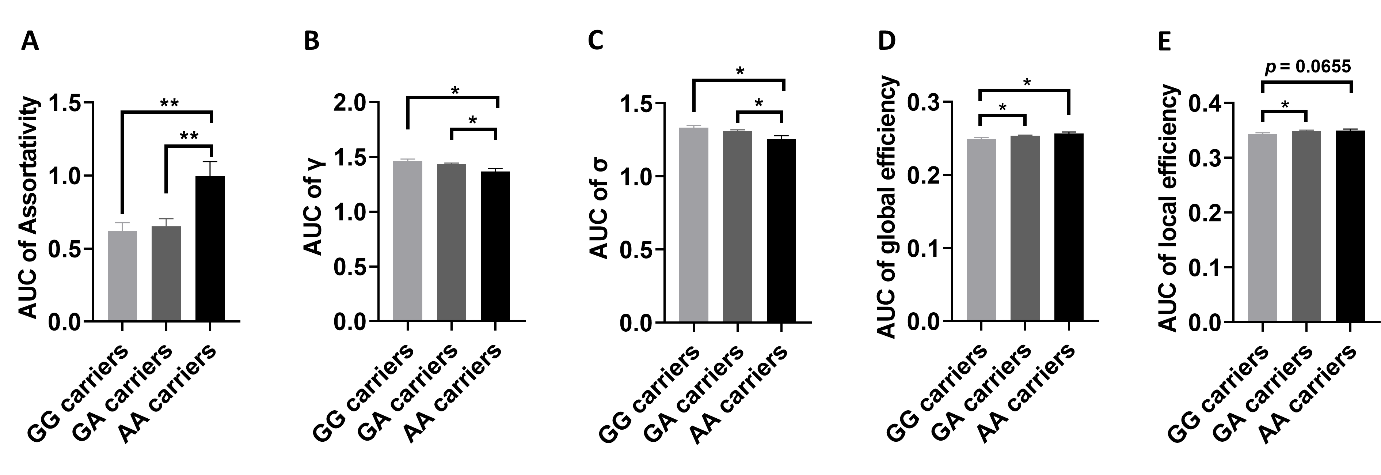

**Figure S1. Group differences of AUCs of global network metrics among GG, GA, and AA carriers.** (A-C) Group differences in AUC of assortativity (A), small-worldness γ (B), and small-worldness σ (C) in structural network. (D-E) Group differences in AUC of global efficiency (D) and local efficiency (E) in functional network. One-way ANOVA test followed by FDR corrections (GG carriers *vs* GA carrier *vs* AA carriers) were used. *p* < 0.05 was considered statistically significant. Abbreviations: AUC, Area under curve; γ, normalized clustering coefficient; σ, small worldness.
